# A national mixed-mode seroprevalence random population-based cohort on SARS-CoV-2 epidemic in France: the socio-epidemiological EpiCov study

**DOI:** 10.1101/2021.02.24.21252316

**Authors:** Josiane Warszawski, Nathalie Bajos, Muriel Barlet, Xavier de Lamballerie, Delphine Rahib, Nathalie Lydié, S Durrleman, Rémy Slama, Rémonie Seng, Philippe Raynaud, Aude Leduc, Guillaume Bagein, Nicolas Paliod, Stéphane Legleye, Cyril Favre-Martinoz, Laura Castell, Patrick Sillard, Laurence Meyer, François Beck, The EPICOV study group

## Abstract

**Background:** the EpiCov study, initiated at the end of the first national lockdown in France, aimed to provide national and regional estimates of the seroprevalence of SARS-CoV-2 infection, and to analyze relations between living conditions and the dynamics of the epidemic. We present and discuss here the survey methodology, and describe the first-round fieldwork.

**Method:** 371,000 individuals aged 15 years or more were randomly selected from the national tax register, stratified by departments, including three overseas departments, and by poverty level with over-representation of people living below the poverty line. Health, socio-economics, migration history, and living conditions were collected through self-computed-assisted web interviews or via computer-assisted telephone interviews. The first-round survey was conducted in May. A random subsample was eligible to receive material for home blood self-sample on dried blood spot (DBS), in order to detect IgG antibodies against the spike protein (Euroimmun ELISA-S), and neutralizing antibodies for non-negative ELISA-S. For the second-round conducted in November, all respondents were eligible for the antibodies detection from home DBS sample, as well as the other household members aged 6 years or more for 20% of them.

**Participation and adjustment for nonresponse:** 134,391 respondents completed the first-round questionnaire from May 2 to June 1, 2020, including 16,970 (12.6%) respondents under the poverty line. Multimodal web/tel interviews was randomly assigned to 20% of the sample. The other were assigned to exclusive CAWI. Overall 17,441 respondents were eligible for home blood sample, among them 12,114 returned the DBS (interquartile date: May 25-June 5). The response probability was first estimated from logit models adjusted on a wide range of auxiliary demographic and socio-economic variables available from the sampling frame, and final weights calibrated to the margins of the population census permitted to correct for a large part of the non-response bias.

**Conclusion:** The Epicov study is one of the largest national random population-based seroprevalence cohort, with both an epidemiological and sociological approaches to evaluate the spread of the COVID-19 epidemic, and the impact on health and living conditions. One of the major interests of this study is the broad coverage of the socio-economic and territorial diversity of the population.

## Introduction

The SARS-CoV-2 pandemic began in Europe at the start of 2020. It immediately generated enormous pressure on the health system, and demands for continual information updates, from public authorities, stakeholders and the general public. In the absence of vaccination and specific treatment options, public health interventions were launched worldwide (1). Each state reacted in its own way, at different speeds and with *ad hoc* solutions based, in particular, on stay-at-home orders, rules for social distancing, the use of personal protective equipment, the isolation of individuals with confirmed infection, the quarantine of their contacts, border restrictions and total or partial lockdowns. The need to follow the evolution of the pandemic, and its influence on living conditions, with sufficient precision and at a fine geographic scale, was common to all countries.

Surveillance systems were set up to estimate the temporal dynamics of the SARS-CoV-2 virus, through monitoring of the number of hospitalizations, deaths, positive virologic and serologic tests in the population, mostly based on data from medical structures, or from repeated cross-sectional studies in various selected populations, such as blood donors or healthcare professionals (2–4). Seroprevalence studies, based on SARS-CoV-2 antibody tests, have been recommended as a means of estimating the cumulative incidence of COVID-19, the disease caused by SARS-CoV-2 (5) Random sampling of the target population remains the gold standard for achieving representativeness (6). However, this approach also requires extensive resources and community engagement for use in the general population, and few SARS-Cov2 seroprevalence surveys based on probability samples have been conducted in general population at national or territorial level (7–12).

The EpiCov study was initiated at the start of the first lockdown in March 2020, to provide an initial point estimate of the seroprevalence of SARS-CoV-2 and a precise description of various socioeconomic and behavioral indicators at the end of the lockdown, at both national and local (*departement*, equivalent to a county) levels. The study was also designed to provide reliable data for the various social groups, including hard-to-reach subgroups, such as people in precarious situations, and to be repeated at different timepoints.

The EpiCov was launched by the National Institute for Health and Medical Research (Inserm), in collaboration with two government Public Statistics offices (Drees and Insee), and the French national public health agency (SPF). It was based on a large random sample of people living in France. The main objectives were to estimate the immunity status both nationally and locally and in various subpopulations, including populations suffering socioeconomic deprivation, to study intrahousehold circulation of the virus, to describe the situation of the population in terms of health status, including mental health, access to healthcare, living conditions (housing, financial status, work, childcare, home-schooling) and interpersonal relationships within the household, including domestic violence, and to study their relations with the serological status.

The first round was performed in May 2020. The second round of the survey took place in November 2020. We describe and discuss here the methodology of the EpiCov cohort.

## Method: design and data collected

### Sampling frame

The sampling frame was the Fideli (Demographic files on dwellings and individuals) (13), a comprehensive database obtained by merging several administrative files, an approach tested several years previously for the French Labor Force Survey (14). These files are updated yearly from annual tax returns, and are curated by the National Institute of Economics and Statistics (Insee), to eliminate duplicates and to identify community housing structures (nursing homes, prisons, military barracks, etc..) and residential hotels for separate treatment. This file covers 96.4% of the population and their dwellings, according to French census data. The Fideli datafile compiled in 2018 was used for EpiCov.

All the dwellings included in Fideli are associated with a mailing address, which can be used to contact the individuals sampled. Additional modes of contact are possible, because at least an e-mail address, landline or mobile phone number was available for 83% of the dwellings in 2018 (one telephone number for at least 69%, one mobile phone number for at least 45%, and one e-mail address for at least 71% in mainland France).

Fideli includes a wide range of relevant auxiliary information at individual and household level. This information is useful for stratification purposes, and for the subsequent correction of non-response bias after data collection.

### Sampling units and target population

The target population consisted of all individuals aged 15 years or older on January 1, 2020, living in mainland France or one of three overseas *departments* (Martinique, Guadeloupe and Réunion Island). Due to the poor quality of the sampling frame, poor internet access and the use of multiple languages, two other overseas *departments*, French Guiana and Mayotte, were excluded from the study.

We also excluded individuals living in prisons at the time of the study, and people living in residential institutions for dependent elderly persons, as caregivers were not available during the epidemic period to help them with internet access or phone calls.

### Sample size calculation

Sample size was calculated so as to ensure sufficient precision for the seroprevalence estimate, the goal being to obtain a 95% confidence interval of 2 points for a prevalence of 5% in administrative subdivisions of 600,000 inhabitants (*department* or *metropolitan area*). This required the collection of at least 500 blood samples from individuals in each of the 96 *departements* in mainland France.

We expected to establish web or telephone contact with 70% (60% in overseas territories) of the initial sample from Fideli, with 70% of these individuals completing the questionnaire (60% in overseas territories), 85% of whom would agree to receive the home sampling kit, with 70% of these individuals mailing the biobank with a dried blood sample, an overall response rate for questionnaires of about 50% for residents of mainland France (35% overseas), and an overall return rate self-samples of 60%.

Based on this calculation, 350,000 individuals were randomly selected from mainland France and 21,000 individuals from the three overseas *departments* included, in order to obtain 170,000 and 7,500 respondents to the questionnaire, and 100,000 and 4,500 tested participants for mainland France and the overseas *departments*, respectively.

### Sampling design

Eligible individuals were selected at random, with stratification by two criteria: administrative area (*departments* in mainland France and three overseas), and a binary indicator of poverty, defined as living over or under a threshold of 60% of the median national *per capita* household income.

Allocations were calculated so as to be proportional to the population of the *department*, but with overrepresentation of the least populated *departments*, to ensure that there were at least 900 respondents in each, assuming a response rate of 50% among those selected. Individuals living in a household below the poverty line were overrepresented in mainland France, constituting 20% of the sample rather than the 13% of the Fideli sampling frame, as a lower response rate was expected for this subpopulation.

The sampling file was also sorted by urban subdivisions, municipality, household income level, and the identification numbers of the dwelling and the individual. This process ensured an implicit stratification for population density, and prevented the selection of two individuals from the same dwelling.

The overall sample was divided into 20 subsamples of 18,550 individuals, called “lots”, separately for French mainland and overseas *departements*, each with the same stratum allocations as for the overall sample. This method was chosen as a flexible means of selecting subsamples to address specific objectives.

The first round of the Epicov survey was performed during lockdown, which limited the supply of materials for kits and laboratory tests. A subsample calculated to obtain about 12,000 tested individuals was planned, including a national subsample of expected 6,000 tests, and five areas displaying overrepresentation, with at least 1,000 expected tests each. Three of these five areas had the highest Covid-19 risk indicators (Haut-Rhin, Paris and the inner suburbs, Oise) and two were considered to be at lower risk (Bas-Rhin and Bouches-du-Rhône) during the first epidemic wave. During the second round of the EpiCov survey (October 26 - December 7, 2020), home sampling was offered to all study participants. Furthermore, for 20% of the participants, this testing included all the other members of their household over the age of six years.

### Data collected: mixed-mode questionnaire and home dry blood samples collected at home

#### Mixed-mode questionnaire

Each of the selected individuals was asked to complete a questionnaire collecting information on several topics. Demographic and socioeconomic characteristics were reported, and health status was described through general and specific questions regarding self-perceived health, symptoms potentially linked to COVID-19, mental health, and access to healthcare, whether or not linked to COVID-19. Questions relating to living conditions were also included: size and nature of the habitation, number of people in the household, etc. Finally, behavior during lockdown was described: work, childcare, home-schooling, inter-partner relationships within the household, etc.

The overall cost of telephone calls and response collection was limited by using two versions of the questionnaire. A short version (taking 26 minutes on average) was proposed for 90% of the sample (18 of the 20 subsample lots). A longer version (34 minutes on average, including all the questions of the short version) was administered to 10% of the EpiCov participants (the two remaining lots).

Both the long and short versions were implemented as self-completed questionnaires, through the computed-assisted web interview (CAWI) system, or were administered by qualified and supervised professional interviews *via* the by computer-assisted telephone interview (CATI) system.

The 370,000 randomly selected individuals were initially sent a personalized contact letter including a presentation of the survey, together with access codes for a web link to the questionnaire. Whenever possible, the information was also provided by e-mail, SMS, and phone call. In all communications, the first name and surname of the selected person from the dwelling were indicated. As the telephone contact details from tax files are not necessarily those of the selected person in the household, the person contacted was asked to forward the letter and notice with the internet link to the intended recipient if necessary.

The availability of telephone interviewers was reduced by lockdown, making it impossible to use CATI for all the subsamples. A concurrent mixed mode was then assigned to three of the 20 subsample lots in mainland France (that is, from the start of the study, interviewers tried to contact selected individuals who had also received the web link to answer online). A sequential mixed-mode was assigned to one subsample lot, in which interviewers tried to reach respondents only two weeks after the start of the study. An exclusive CAWI mode was assigned to the other 16 subsamples. The number of CATI lots was higher for the overseas departments.

Non-responses were minimized, by sending multiple postal, e-mail, SMS, and vocal messages to participants, to remind them of the importance of their participation in EpiCov.

#### Home sample collection kits

Serological tests was based on a capillarous blood samples collected by the participants themselves, at home, from a finger prick dried blood spot 903 Whatman paper (DBS) kits, which was subsequently analyzed in a centralized virology laboratory.

The home sample was proposed during the telephone or online questionnaire, with appropriate explanations about how the test should be performed. Respondents were informed that these serological tests were destined for research purposes, and that the results would only be returned to the individual much later.

The sample kits were delivered by express mail to each participant agreeing to undergo testing. The kits included all the necessary material and printed instructions on how to perform the sample, together with a pre-paid addressed envelope for the return of the dried-blood sample to the EpiCov biobank (CRB-Centre Hospitalier Universitaire Pellegrin, Bordeaux, France). A hotline telephone number was provided to allow participants to pose any questions they might have.

At the biobank, DBS cards were first store in 2D FluidX 96-Format 0.5 mL tubes (Brooks) at −30°C. Once most of the cards were received, four 4.7 mm discs were punched from the spots on the Whatman paper, using a PantheraTM machine (PerkinElmer).

The tubes were sent to the virology laboratory (Unité des virus Emergents, Inserm/IRD, Marseille, France) for elution of the dried blood and serological analysis. Eluates were processed with a commercial ELISA (Euroimmun®, Lübeck, Germany) to detect anti-SARS-CoV-2 antibodies (IgG) directed against the S1 domain of the spike protein of the virus (ELISA-S), according the manufacturer’s instructions. All samples with an ELISA-S test optical density ratio ≥ 0.7 were also tested with an in-house micro-neutralization assay to detect neutralizing anti-SARS-CoV-2 antibodies (SN) (15). For these tests, VeroE6 cells cultured in 96-well microplates, 100 TCID50 of SARS-CoV-2 strain BavPat1 (courtesy of Prof. Drosten, Berlin, Germany) and serial dilutions of serum (1/20–1/160) were used. Dilutions associated with the presence or absence of a cytopathic effect on post-infection day 4.5 were considered to be negative and positive, respectively.

## Ethics and regulatory approval

The serological results were sent to the participants by post, at the end of the study, with advice concerning the lack of scientific knowledge about individual protection against future re-infection for those testing positive for antibodies.

The study protocol was approved by the appropriate committees. The EpiCov cohort protocol was approved by the CPP (“*Comité de Protection des Personnes”*, French equivalent of the Research Ethics Committee) on April 24, 2020, and amended on June 5. A notice of opportunity from the National Council for Statistical Information (CNIS) was obtained on April 17, 2020, and approval was obtained from the *Comite du label de la statistique publique* on April 21, 2020, proving its adequacy to statistical quality standards, and from the CNIL *(“Commission nationale de l’informatique et des libertés”*, the French independent administrative authority responsible for data protection) on April 25, 2020. All these committees modified their procedures to ensure rapid processing and a fast implementation of the study.

## Fieldwork

The first round of the EpiCov cohort study began on May 2nd, 2020, and questionnaires were collected until June 1, 2020.

The telephone numbers available in the Fideli sampling frame were supplemented by a telephone directory search, which increased the proportion of available numbers from 71% to 81%. A letter was mailed to 370,928 (349,936 participants in mainland France and 20,992 in the overseas *departments*. As postal services were not fully functional in France during this period, e-mails and SMS were also sent, at the same time when possible: 258,867 e-mails (246,019 in mainland France and 12,848 in the overseas *departments*) and 165,028 SMS. Only 4.2% of the letters remained undistributed, 7.4% of the e-mails and 17% of the SMS were bounced back as spam. Interviewers reported that some respondents contacted by telephone had not received the initial letter in time, almost certainly due to poor mail distribution during the pandemic period or because they had left their usual place of residence during the lockdown period.

Sequential reminders were sent with different formulations and modalities (Supplementary Fig S1): 253,801 letters, 163,434 and 148,820 e-mails at two different times, 116,600 and 97,505 SMS, 112,578 voice messages on mobile phones and 26-856 on landline telephones. Each reminder conducted to a new peak in completed questionnaire (Figure 2).

**Figure 1.**
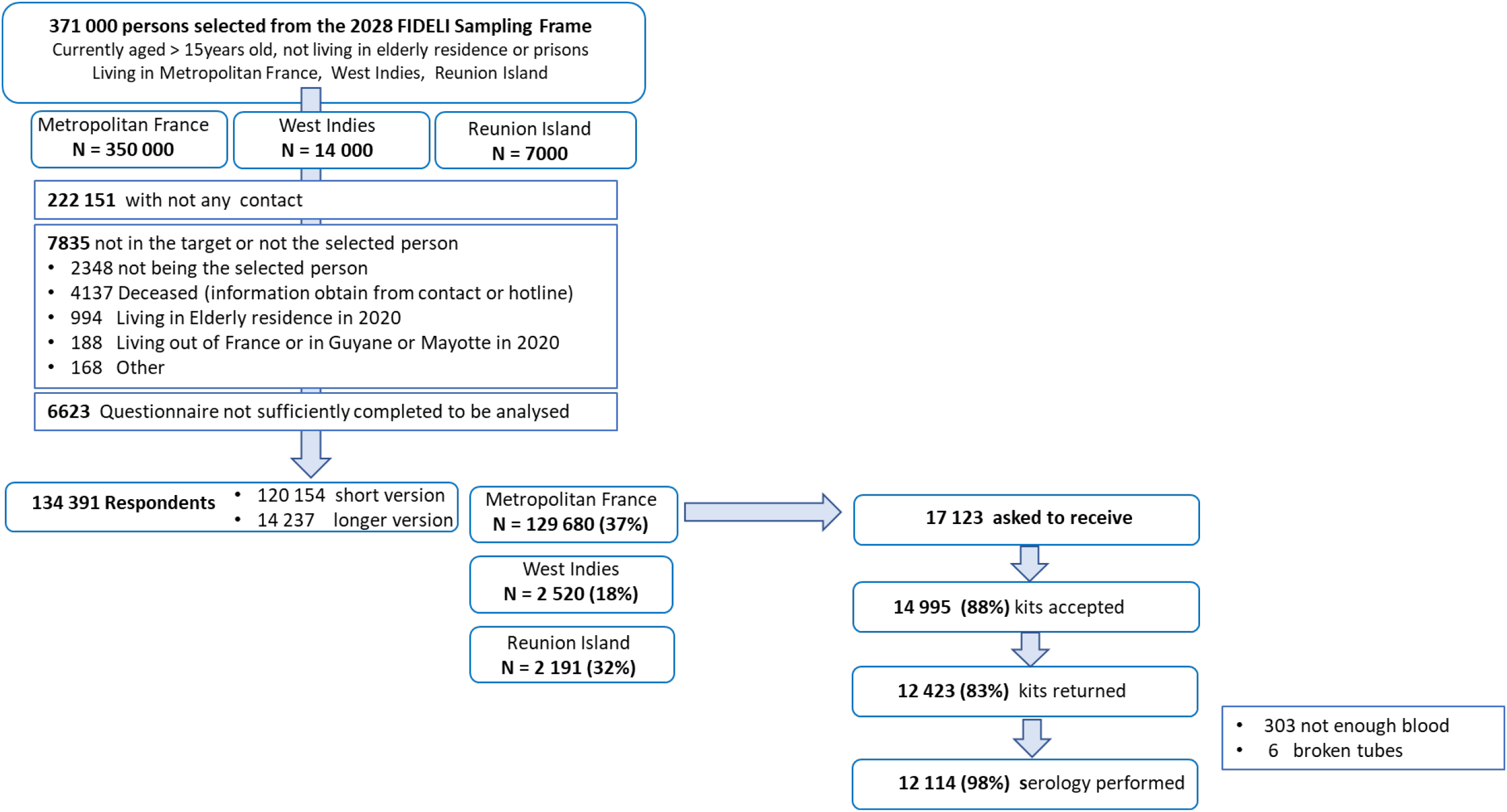
Flow cha rt. The EpiCov study - Round 1, May 2020.

**Figure 2.**
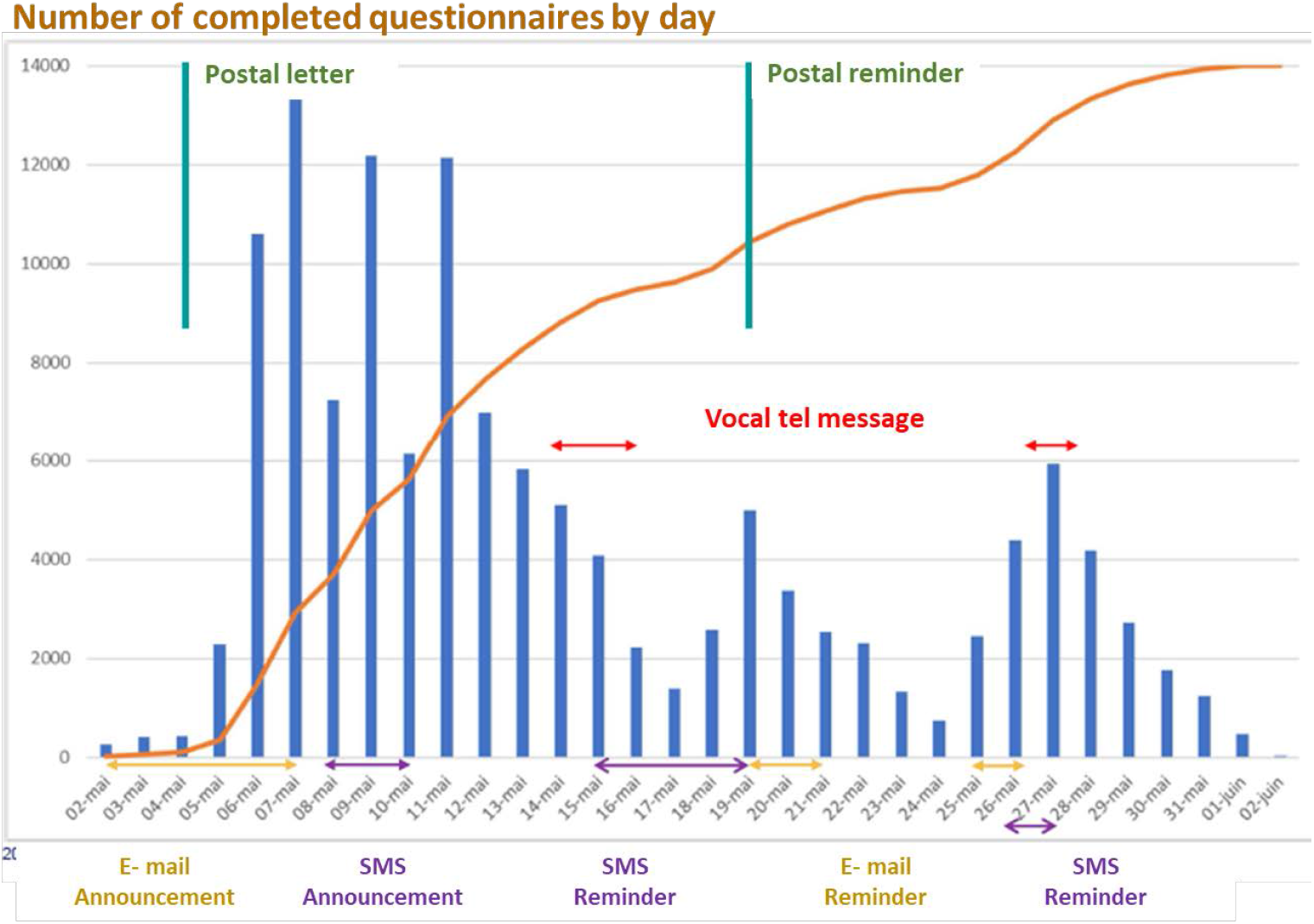
Change in the number of interviews over time, as a function of the sending of reminders. **The EpiCov study - Round 1, May 2020**

The home sampling kits were sent to eligible participants who agreed to home testing, from May 7 to June 4. By May 21, 50% of the participants who had agreed to receive a kit had already drawn their blood sample (IQR: 18-28); and the dry-spot samples were returned to the biobank between May 15 and August 3, with a median return date of May 29 (interquartile: May 25 - June 5) (Figure 3). Overall, 11,882 reminders to use the blood kit were sent, from May 20 to June 26: 76% by SMS, 21% by e-mail, and 3% by voicemail. Difficulties with the sampling process reported to the hotline led to 217 kits being resent.

**Figure 3.**
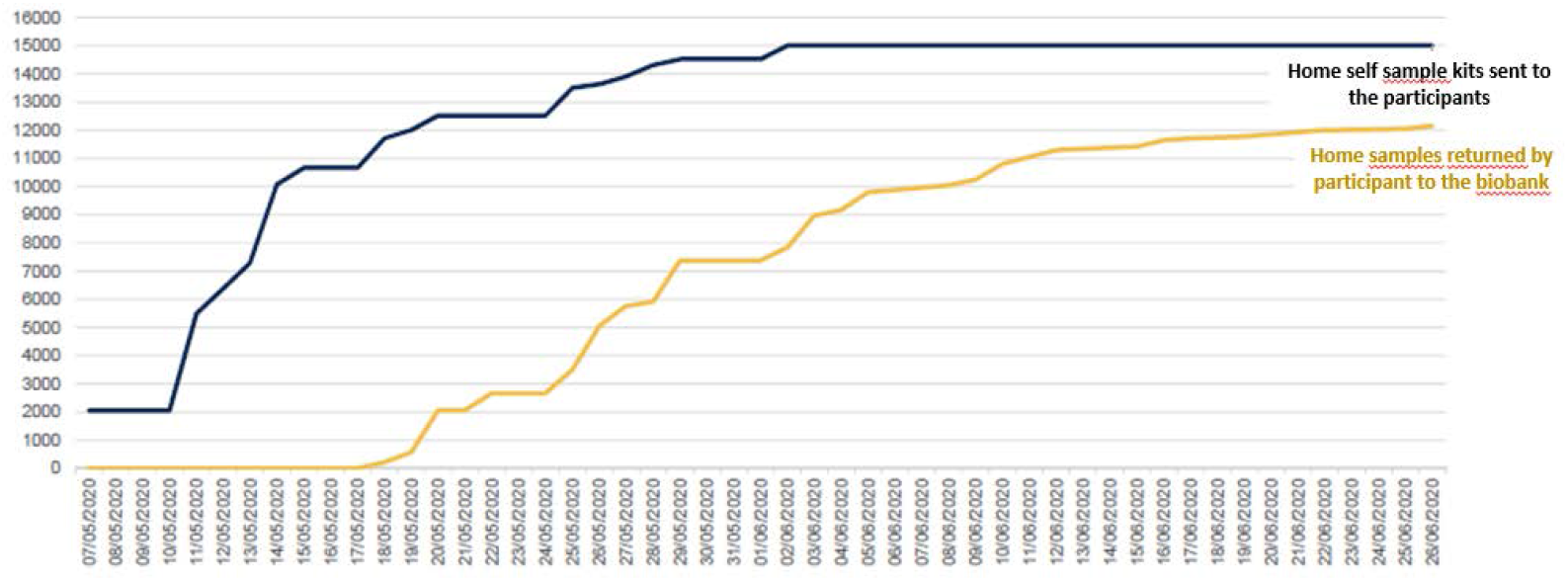
Number of home sampling testing kits accepted and number of samples returned to the biobank center over time. **The EpiCov study - Round 1, May 2020**

## Participation rate (flow chart in figure 1)

No contact was established for 222,151 of the individuals selected. For the others, a specific assessment was performed to check that the respondent was indeed the selected individual, by comparing the sex and birth date recorded on the questionnaire with those available from the Fideli sampling frame. This assessment led to the exclusion of 2,348 individuals who were not the selected individual. Another 5,487 individuals were considered to be outside the sampling frame (deceased, living in a communal institution for the elderly, no longer living in France, or living in French Guiana or Mayotte), and 6,623 completed too few items for retention in the final database.

There were 134,391 respondents in total (Fig. 3): 120,154 completed the short form of the questionnaire and 14,237 completed the long questionnaire. The response rate was 37% in mainland France, 32% for Réunion Island and 19% in the French West Indies. For the multimode subsample lots, contacted by internet and telephone, the response rate was 46% in mainland France, 41% for Réunion Island and 33% in the French West Indies (Supplementary Tab. 1). For those contacted solely by the Internet, the response rate was 35% in mainland France, 24% in Reunion Island and 23% in the French West Indies. For those responding via the Internet, 20% did so on a smartphone and 7% on a computer tablet. In mainland France, the response rate for people living below the poverty threshold was lower than that for the population as a whole, by a factor of 1.7.

In total 17,123 respondents were eligible for home testing and asked to receive a kit, of whom 14,995 (88%) agreed to receive the kit, and 12,423 (83%) returned a sample. In total, 12,114 tubes were analyzed, after excluding 303 kits with too little blood and 6 tubes broken at the laboratory, corresponding to a tested rate of 71% of the total eligible subsample respondents.

## Non-response adjustment weights

Three sets of final calibrated adjusted weights were produced to correct for non-response: for the whole respondents who completed the common short questionnaire, for the 10% subsample who completed the extended version of the questionnaire, and for the subsample who returned the home sample to biobank for serological test.

The method was similar in each case. In the first step, the survey weight (the inverse of the inclusion probability) was divided by an estimate of the probability of response. Response probabilities were estimated using logit models adjusted for auxiliary variables potentially linked to both the response mechanism and the main variables of interest in the EpiCov survey. The sampling frame provided many rich auxiliary demographic and socioeconomic variables, 90 of which were correlated with at least one variable of interest in at least one *department*. These variables also included the quality of contact information, and variables aggregated at local area of the place of residence, such as population density, proportion of people aged over 65 years or below the poverty line, obtained from geo-referencing information including in the Fideli sampling frame. Response homogeneity groups were derived from this estimated probability, established within each department for correction of non-response correction to the common short questionnaire sample. The response probability was then estimated from the percentage of respondents in each homogeneity group, yielding first-step weights. The weight of a respondent was as high as the response rate was lower in the homogeneity group it belongs, resulting in a correction of the effects of differences in response rates between subgroups.

In the second step, these weights were calibrated according to the margins of the population census data and population projections for several variables (10-year age categories, by gender, *departement*, and diploma level, by region). Weights for the serological subsample were calibrated at national and local level for the five overrepresented areas. This calculation was designed to decrease the variance and the residual bias for variables correlated with margins.

We found that there was an endogenous selection effect concerning the declaration of COVID-19-associated symptoms, as CAWI-respondents reported more symptoms than CATI-respondents. We accounted for this, by using a Heckman model (16–18) to generate specific weights to correct for this bias in the estimation of symptom prevalence. This estimate was based on the simultaneous modeling of participation and output variables (collected by the survey). For identification purposes, the model needs an instrument variable, explaining survey participation but not playing any role in the output variables. In the case of EpiCov, this instrument was the binary variable distinguishing between people selected from the mixed CAWI/ CATI subsample lots and those in the pure CAWI subsample (Supplementary Table S2) (19). This binary variable was a reliable instrument, because the 20 subsample lots derived from a random splitting of the whole sample, whereas the participation rate was about 10 points higher in the four CAWI/CATI lots than in the 16 pure CAWI lots (Supplementary Table S3). In the general weight estimation framework presented above, the Heckman step replaces the logit model step, in case endogenous selection is detected, everything else remaining equal.

## Discussion

### Strengths

The Epicov cohort is one of the largest national representative population-based seroprevalence surveys of individuals aged 15 years and over, using both an epidemiological and sociological approaches to evaluate the spread of the emerging COVID-19 epidemic and its impact on health and living conditions. One of the major interests of this study is the broad coverage of the socio-economic and territorial diversity of the population, including three overseas French departments.

Epicov was initiated at the end of March, just after the start of the first national lockdown in France. It was implemented rapidly, during an extremely challenging period, to provide a first point estimate of the situation at the end of the lockdown, corresponding to the end of the first wave of the COVID-19 epidemic. This initial information will be invaluable for comparisons with the ongoing second round of the EpiCov study, corresponding to the second lockdown in France to deal with a new peak in the epidemic emerging in the fall of 2020. Only two other national serological studies based on random samples of general population were reported to be implemented before 2020 summer, in Spain (20) and in England (10).

As we overrepresented people living under the poverty line, which permitted to include 17 000 of them, and as many information on socio-economic and first and second generation of migration status were available in the sampling frame and/or collected through the questionnaire, it is possible to focus on social inequalities in the context of the epidemic crisis with powerful specific analysis.

The Fideli administrative database as a sampling frame had many advantages. It provides a very high coverage of the target population, with postal addresses for all, and additional e-mail addresses or telephone numbers for most, and a rich set of auxiliary information useful for non-response correction. A mixed-mode web/telephone data collection approach was decided early in the project, to limit selection bias which should have occurred with a single collection mode, because internet and telephone access are not uniformly distributed between age groups, areas of residence and educational levels. This decision was consistent with the general guidelines for social surveys (17,18). A recent meta-analysis on a corpus of 45 mixed-mode and 51 single-mode surveys (23) showed that mixed-mode surveys were of better quality, in terms of representativeness, than single-mode surveys. To a certain extent, the transition to mixed-mode surveys (20), including online data collection, was already underway worldwide, but has been accelerated by the pandemic (24).

We selected directly individuals from the sampling frame, rather using first-stage household units. Both options were possible with Fideli, but the former, though more complex to implement, had the advantage of avoiding overrepresentation of the households’ tax registrants. Tax registrants are inconsistent in the provision of e-mail addresses or phone numbers for the other members of their households, and are likely to be the member of the household with the highest income or social position. Some individuals tried to respond in place of the selected person, as observed for 2348 contacts. We then anticipated a larger proportion of non-responses than with household sampling, because the first person contacted may refuse or forget to transmit the announce letter or email to the eligible one, particularly if not at home during the survey.

The maximum recommended duration for telephone surveys is half an hour (25,26), and 20 minutes for web surveys (27), with a median time of 10 minutes being considered ideal (28). Needed by the multiple objectives of EpiCov, the common short questionnaire administered to 90% of the participants slightly exceeded these duration (25-minute average). A longer version for the remaining 10% permitted to explore living conditions and mental health in more detail. Less than 5% of the questionnaires were insufficiently completed to be retained for analysis, and there were quite few missing values for the others.

Home self-sampling for the detection of SARS CoV-2 antibodies was an innovative process. In the context of the lockdown, the epidemic peak, and the low availability of surgical masks, it did not seem feasible to send nurses to the homes of all the participants or to ask the selected participants from all over France to go to medical analysis laboratories for testing. Home sampling appeared to be the best method for limiting self-selection bias. The acceptability of self-sampling based on dried-blood spots had already been demonstrated in 2016, for a study of the seroprevalence of HIV and hepatitis B & C based on a random telephone survey in the general population in France (29). However, this approach had never been used for the detection of coronavirus antibodies. The Euroimmun ELISA-S test chosen for this study with ratio ≥ 1.1 has a sensitivity of 94.4%, according to the manufacturer. It was evaluated in different studies showing a specificity ranging from 96.2% to 100% and sensitivity ranging from 86.4% to 100% (30,31,31,32). The neutralization assay, considered as the most specific serological assay capable of detecting true positive cases and functional neutralizing antibodies, for the highest dilution of serum yielding a positive result, had a specificity close to 100% with titers ≥40 considered positive in a population of blood donors sampled in 2017-2018 (15).

As a very large number of auxiliary demographic and socio-economical variables were available from the sampling frame used for EpicCov, it was possible to produce accurate estimation of the response probability from logit models, and then calibrated adjusted weights able to correct a large part of the non-response bias. Non-response is a crucial issue to ensure the representativeness of the EpiCov study, the major objective of which was to provide descriptive indicators for people living in France, including estimates of the prevalence of antibodies against SARS-Cov-2, the prevalence of symptoms, and the prevalence of various living condition indicators during lockdown.

Moreoever, EpiCov provided us an exceptional opportunity to correct for endogenous self-selection bias, which concerns studies dealing with a topic of major impact on the population, and considerable media coverage. In the context of the Covid-19 epidemy, people feeling most concerned about the disease, whether worried, personally affected or related to patients who had suffered the disease, would be more likely to participate than others. Such self-selection biases have been largely described (33,34), particularly in the context of health crises (35). The leverage-saliency theory (33) suggests that the decision to respond to a survey depends mostly on concern about the topic of the survey and the response burden, in addition to the sponsor’s notoriety and status. This is particularly likely when self-interview modes are used, and less likely when interviews are conducted by phone or, more efficiently, face-to-face. It is likely that such endogeneous bias may conduct to overestimate the proportion of symptomatic infection in most reported studies. Classical non-response models cannot deal with such endogenous bias. In the EpiCov study, the multimodal random subsample constituted a reliable instrument to perform the Heckman model (16–18), conducted to specific weights to correct overestimation in the proportion of reported symptoms. This correction was made at the expense of a higher variance. However, this method has demonstrated the magnitude of the problem, too often neglected, as it cannot be neither evaluated and corrected in most all studies.

## Limitations

We did not included people living in residences for elderly. It was not feasible during the first lockdown. Specific studies will be required to provide reliable estimates for these populations, greatly affected by the epidemic.

In recent decades, willingness to participate in population-based surveys has decreased considerably worldwide (36,37). We anticipated an overall response rate at about 50% in mainland France, including 70% of initial web or telephone contact, among them 70% of completed questionnaires. This took into account the difficulties to reach the selected individual in a household as explained above, and relocations and deaths since 2018-last updated date of the Fideli sampling frame.

Calculation for the sample selection was performed considering multimodal web/telephone data collection for the whole sample. However, due to the limited number of simultaneous telephone interviewers during lockdown, we decided to offer multimodal interview for only four of the 20 random subsample lots (extended to five in the French West Indies and nine on Reunion Island), giving an overall response rate of 36%. It should be stressed that the response rate for the multimodal lots was close to that expected: 45% for the three concurrent mixed-mode subsamples and 44% for the sequential mixed-mode subsample. The CATI response rate reached 46% in concurrent multimodal administration, but did not exceed 29% for sequential multimodal administration, confirming that the sequential approach optimized the use of the Internet without decreasing the overall response rate, consistent with the general recommendation of such approaches (21–23).

The response rate was also affected by the short duration of the fieldwork, limited to four weeks centered on the end of the lockdown (May 11). Each reminder led to a response rate peak, even the last reminder sent five days before the end of the study, suggesting that it remained a potential for the response to increase over time.

The lower-than-expected rate of response to the survey did not decrease the statistical power for the main stringent serological outcomes, as it was counterbalanced by the higher-than-expected rate of acceptance of home sample, with 88% acceptance and 83% sample return rates, versus the expected levels of 85% and 70%, respectively planned. The epidemic context, the reputations of well-known public institutions mentioned regularly in the media during this period, the association with public health researchers pointed out in the information leaflet, and the possibility of receiving the serological result probably contributed to the high return rates. At the time, very few virological and serological tests were available in France, and many people were interested in knowing whether they had been infected.

## Conclusion

The EpiCov cohort study provides a reliable epidemiological and sociological tool for improving our understanding of the spread of SARS-CoV-2 and its multiple societal consequences. It improves assessments of the efficacy of public health measures, and provide policy-makers with support for the management of the epidemic on the basis of scientific evidence.

## Data Availability

The datasets used and analyzed during the
current study are available from the corresponding author on reasonable request.

## Acknowledgements

The authors first sincerely thank all the participants to the EpiCoV study.

We thank greatly the INSERM staff, and especially Karim Ammour, Jean-Marc Boivent, Sophie Circosta, Jean-Marie Gagliolo, Michael Hisbergues, Frédérique Le Saulnier, and Frédéric Robergeau,. who participated with an exceptional engagement to make it possible to develop, in a particularly short period of time, and to maintain all regulatory, budgetary, technical, and logistical aspects of the EpiCov study.

We thank greatly the staff of Santé Publique France, and especially Lucy Duchesne, who played a major role in the organization and quality of the seroprevalence component of the EpiCov study.

We thank greatly the CRB biobank staff (Centre Hospitalier Universitaire Pellegrin, Bordeaux, France), and especially her head, Dr Isabelle Pellegrin, for the quality of the management process of DBS samples, implemented in such a short period fort the first-round of the EpiCov study.

We thank greatly the UVE virology department staff, and especially Toscane Fourié, for the high quality in management of such a large amount of serological assays.

We thank greatly the Ipsos staff, and especially Christophe David and Valérie Blineau, which highly contributed to the quality of data collection.

## SUPPLEMENTARY TABLES AND FIGUES

**Figure S1.**
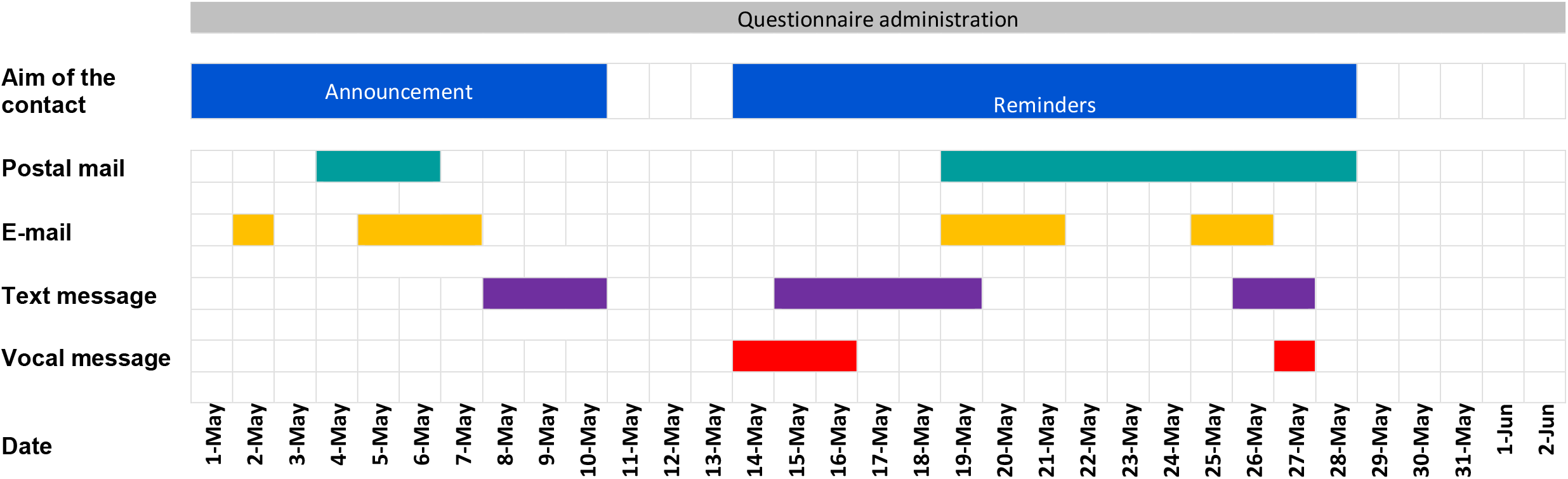
Reminders timetable – EpiCov Round 1 – May 2020.

**Table S1.** Response rates according to the type of contacts for the people selected. The EpiCov study - Round 1, May 2020.

| Initial contact details available | Mainland France<br>N=350 000 |  |  |  | West Indies<br>N=14 000 |  |  |  | Réunion Island<br>N=7 000 |  |  |  |
| --- | --- | --- | --- | --- | --- | --- | --- | --- | --- | --- | --- | --- |
|  | Initial frame | Enriched | Response rate |  | N=14000 | Enriched | Response rate |  | Réunion<br>N=7000 | Enriched | Response rate |  |
|  | N(%) | N(%) | n | % | N(%) | N(%) | n | % | N(%) | N(%) | n | % |
| Postal address (all) | 349 935 |  | 129 507 | **37%** | 13994 |  | 2 645 | **19%** | 6 998 |  | 2 239 | **32%** |
| E-mail only | 40 466 (12%) | 35 699 (10%) | 11 124 | **31%** | 1 569 (11%) | 1432 (10%) | 178 | **12%** | 877 (13%) | 658 (9%) | 147 | **22%** |
| Phone number only | 40 309 (12%) | 46 961 (13%) | 9282 | **20%** | 1 952 (14%) | 2254 (16%) | 327 | **15%** | 839 (12%) | 1 340 (19%) | 356 | **27%** |
| E-mail + phone | 209 382 (60%) | 210 350 (60%) | 102 211 | **49%** | 7 083 (51%) | 7097 (51%) | 1983 | **28%** | 3 538 (51%) | 3 661 (52%) | 1 599 | **44%** |
| No mail or phone | 59 766 (17%) | 59 956 (16%) | 6890 | **12%** | 3 390 (25%) | 3211 (23%) | 157 | **5%** | 1 744 (25%) | 1 339 (19%) | 137 | **10%** |

**Table S2.** Assignment to random subsample lots – The EpiCov study, Round 1 – May 2020

|  | Contact mode |  |  | Questionnaire version | Home sampling kit proposition |  |  |  |  |  |  |
| --- | --- | --- | --- | --- | --- | --- | --- | --- | --- | --- | --- |
| Lot | Mainland France | West Indies | Réunion Island | Short / Long | National sample | Haut Rhin | Paris | 92-93-94 | Bas-Rhin | Oise | Bouches du Rhône |
| 1 | Multimode | Multimode | Multimode | Long | No | Yes | Yes | Yes | Yes | Yes | Yes |
| 2 | Multimode | Multimode | Multimode | Short | Yes | Yes | Yes | Yes | Yes | Yes | Yes |
| 3 | Multimode | Multimode | Multimode | Short | No | Yes | Yes | Yes | Yes | Yes | Yes |
| 4 | Multimode | Multimode | Multimode | Short | No | Yes | Yes | No | Yes | Yes | Yes |
| 5 | Pure Internet | Multimode | Multimode | Short | No | Yes | Yes | No | Yes | Yes | Yes |
| 6 | Pure Internet | Pure Internet | Multimode | Short | No | Yes | Yes | No | Yes | Yes | Yes |
| 7 | Pure Internet | Pure Internet | Multimode | Short | No | Yes | No | No | Yes | Yes | Yes |
| 8 | Pure Internet | Pure Internet | Multimode | Short | No | Yes | No | No | Yes | Yes | Yes |
| 9 | Pure Internet | Pure Internet | Multimode | Short | No | Yes | No | No | Yes | No | Yes |
| 10 | Pure Internet | Pure Internet | Pure Internet | Short | No | Yes | No | No | Yes | No | Yes |
| 11 | Pure Internet | Pure Internet | Pure Internet | Short | No | Yes | No | No | Yes | No | Yes |
| 12 | Pure Internet | Pure Internet | Pure Internet | Short | No | Yes | No | No | Yes | No | Yes |
| 13 | Pure Internet | Pure Internet | Pure Internet | Short | No | Yes | No | No | Yes | No | No |
| 14 | Pure Internet | Pure Internet | Pure Internet | Short | No | Yes | No | No | Yes | No | No |
| 15 | Pure Internet | Pure Internet | Pure Internet | Short | No | Yes | No | No | No | No | No |
| 16 | Pure Internet | Pure Internet | Pure Internet | Short | No | Yes | No | No | No | No | No |
| 17 | Pure Internet | Pure Internet | Pure Internet | Short | No | No | No | No | No | No | No |
| 18 | Pure Internet | Pure Internet | Pure Internet | Short | No | No | No | No | No | No | No |
| 19 | Pure Internet | Pure Internet | Pure Internet | Short | No | No | No | No | No | No | No |
| 20 | Pure Internet | Pure Internet | Pure Internet | Long | No | No | No | No | No | No | No |
| **Number of concerned lots concerned** | **4** | **5** | **9** | **2** | **1** | **16** | **6** | **3** | **14** | **8** | **12** |

**Table S3.** Response rate according subsample lots. The EpiCov study - Round 1, May 2020.

|  | Mainland France |  |  | French West Indies |  |  | Réunion Island |  |  |
| --- | --- | --- | --- | --- | --- | --- | --- | --- | --- |
|  | *N* | *n* | % | *N* | *n* | % | *N* | *n* | % |
| **TYPE OF RANDOM SUBSAMPLE LOT** | **Selected** | **Lots** |  | **Selected** | **Lots** |  | **Selected** | **Lots** |  |
| Single mode (web only) | 279 954 | 16 |  | 9 096 | 13 |  | 3 848 | 11 |  |
| Tel/web multimode | 69 982 | 4 |  | 4 898 | 7 |  | 3 150 | 9 |  |
| **RESPONSE RATE** | **Selected** | **Included** |  | **Selected** | **Included** |  | **Selected** | **Included** |  |
| **Overall** | 350 000 | 129 507 | 37% | 14 000 | 2 645 | 19% | 7 000 | 2 239 | 32% |
| **By type of subsample lot** |  |  |  |  |  |  |  |  |  |
| Single mode (web only) | 279 954 | 97 524 | 35% | 9 096 | 2 053 | 23% | 3 848 | 941 | 24% |
| Tel/web multimode | 69 982 | 31 983 | 46% | 4 898 | 1 593 | 33% | 3 150 | 1 298 | 41% |
| **By poverty status stratum** |  |  |  |  |  |  |  |  |  |
| **All types of lot** |  |  |  |  |  |  |  |  |  |
| Below the poverty line | 69 997 | 16 970 | 24% |  |  |  |  |  |  |
| At or above the poverty line | 279 939 | 112 537 | 40% |  |  |  |  |  |  |
| **Single mode (web only)** |  |  |  |  |  |  |  |  |  |
| Below the poverty line | 55 998 | 12 513 | 22% |  |  |  |  |  |  |
| At or above the poverty line | 223 956 | 85 011 | 38% |  |  |  |  |  |  |
| **Single mode (web only)** |  |  |  |  |  |  |  |  |  |
| Below the poverty line | 13 999 | 4 457 | 32% |  |  |  |  |  |  |
| At or above the | 55 983 | 27 526 | 49% |  |  |  |  |  |  |
| **PROPORTION WITH LONG VERSION** | **Included** | **Short version** |  | **Included** | **Short version** |  | **Included** | **Short version** |  |
| **All** | 129 507 | 13 933 | 11% | 2 645 | 299 | 11% | 2 239 | 231 | 10% |
| **By mode of interview** |  |  |  |  |  |  |  |  |  |
| Web interview | 117 579 | 10 371 | 9% | 1 524 | 141 | 9% | 1 409 | 140 | 10% |
| Tel interview | 11 928 | 3 562 | 30% | 1 121 | 158 | 14% | 830 | 91 | 11% |

